# Breaking Free from Tobacco: Insights from Chipangali’s Small-Scale Farmers on Embracing Alternative Crops in Zambia

**DOI:** 10.1101/2025.02.20.25322609

**Authors:** Mercy Mataliro, Martha Mutalange, Wingston Felix Ngambi, Cosmas Zyambo

## Abstract

Tobacco is an important cash crop for farmers across the globe but has substantial health and environmental risks, thus the World Health Organization Framework Convention on Tobacco Control (WHO FCTC) emphases the need for countries to promote alternative crop farming. This study seeks to understand the success of the alternative crop promotion program among tobacco small-scale farmers and government officials in Chipangali district, Zambia. The study analyzed data from qualitative key-informant interviews (KIIs) with government officials and extension officers (n = 5) and focus group discussions (FGDs) with farmers (n = 6). The FGDs and KIIs were audio recorded, transcribed, and then translated into English. An inductive thematic analysis was conducted to identify challenges and lessons learned from alternative crop farming program provided by participants. We highlight important factors linked to the success of the alternative crop promotion program, challenges and lessons learned. The results highlight several factors, including access to farming inputs, market dynamics, government support and farmers’ perceptions of tobacco farming associated with success of the program. Challenges identified included high faming input costs, market uncertainties, limited support, and dependency on tobacco companies. The alternative crop promotion program provided valuable lessons to farmers. However, majority of farmers felt the program was not inclusive and such initiative in future should accommodate all farmers to be successful. Additionally, provision of farming inputs and improvement of supply chain for the promoted crops could motivate tobacco farmers to transition to alternative crop cultivation. Furthermore, addressing environmental challenges such as water scarcity and unpredictable weather patterns through adaptive measures like drought-resistant crops and irrigation systems, could contribute to the success of the program. This study highlights the concerns and lessons that tobacco farmers raised which could be incorporated in the implementation of alternative crop promotion program in Chipangali.

## 1. Introduction

Despite the World Health Organization Framework Convention on Tobacco Control (WHO FCTC) articles 17 and 18 specifically addressing environmental concerns related to tobacco cultivation and promotion of transition to more sustainable practices such as alternative crop farming (1, 2). Studies in Indonesia, the Philippines, Malawi, Kenya, and Zambia show that most farmers venture into tobacco farming due to the perceived economic viability of tobacco (3–6). Due to the continued production of tobacco, 8.71 million deaths and 229.8 million disability-adjusted life years occur worldwide (7, 8). In many low- and middle-income countries (LMICs), agriculture is an important economic sector, contributing significantly to GDP and providing employment for a large proportion of the population (9–11). Cash crops such as tobacco in particular are considered important drivers of development and employment (12) especially in LMICs (13). However, tobacco farming poses substantial health and environmental risks (14, 15).Tobacco cultivation is associated with degradation of ecosystems and natural resources, habitat loss, livestock deaths, soil erosion, water and air pollution (16).

Given the health and environmental risks associated with tobacco cultivation, tobacco farming is facing increasing regulation. Thus, several Governments and policymakers across the world are urged to support tobacco farmers in transitioning to alternative crop farming (17). However, tobacco control in tobacco-growing countries especially sub-Saharan Africa faces significant obstacles due to economic arguments and conflicting priorities within politically powerful ministries such as trade, finance, agriculture, and labour (18). These ministries often prioritize economic growth and employment, which are heavily supported by the tobacco industry (1). As a result, efforts to reduce tobacco cultivation are perceived as threats to economic stability and livelihoods, leading to resistance against tobacco control policies (19).

In Zambia, the epidemiological studies in both adolescents and adults have shown a steady trend of between 8.7% -12.3% with the rural and males smoking more than the urban and the females respectively(20, 21). Zambia is among the top 5 tobacco-growing countries in Africa (22). In the 2021/22 season alone, a total of 21,612 tobacco farmers were registered in Zambia and cultivated about 19,747.90 hectares of land compared to the previous record for the number of registered tobacco farmers of about 18,000 in the 2020/2021 season, this represents an increase of about 20.1% in the number of registered farmers. For the 2022/2023 farming season, Zambia registered approximately 23,000 tobacco farmers, this represents an increase from the 21,612 farmers registered in the 2021/2022 season approximately 6.42% increase (23), this highlights the major challenge of switching to tobacco farmers to alternative crops. This increase in the high number of farmers registering to grow tobacco in Zambia underscores the need for evaluation of alternative crop implementation programs.

To our knowledge, there is a lack of in-depth analyses exploring the promotion of alternative crop programs cultivation by Small Scale Tobacco Farmers in Zambia. This study aims to bridge that gap by exploring the challenges of alternative crop cultivation promotion program and draw lessons leant from the program. Notably, Zambia currently lacks a structured tobacco growing policy for farmers who want to go into alternative crops. The findings from this study can inform the development of targeted alternative crop interventions, which are critical for reducing amounts of tobacco grown in the Zambia hence reducing the burden of non-communicable diseases (NCDs) and improving overall health outcomes.

## 2. Materials and Methods

This study employed a qualitative description approach, a methodology best suited for problem identification, hypothesis generation, theory formation and concept development (24). Qualitative methods highlight the perspectives of those interviewed, allowing researchers to identify and describe themes and patterns across individual perspectives. Data was collected using focus group discussions (FGDs) and semi-structured key informant interviews between January and February 2025. The two sources of data allowed us to gain an insight into the perspectives of the alternative crop promotion program among tobacco farmers who participated and those who did not in Chipangali district. The FGDs were used to explore farmers perspectives about the alternative crop promotion program. A total of six (6) FGDs consisting of 10 participants were conducted among tobacco farmers in Chipangali district. Three FGDs were conducted among tobacco farmers who participated in the alternative crop promotion program and the other three FGDs with those who did not. Selection of participants for the focus groups discussions was done using purposive sampling. The focus of the FGDs was to explore the challenges and barriers faced by tobacco small-scale farmers in transitioning to alternative crops. The FGD was chosen because it allows one to obtain a wide range of perspectives and insights on a particular subject topic, fostering exchanges among participants and stimulating debate leading to in-depth information and insights compared to an individual interview. It is also useful in situations when participants share common practices in a shared context (25), in this case growing tobacco and being involved in alternative crop program. The FGDs were conducted in Chichewa which is the local language of the study area. Key informant interviews were conducted over a two-week period in January 2025 in the key tobacco-producing areas of Chipangali district of Zambia. Informants included five (n = 5) government officials in Chipangali district. These informants occupied positions such as provincial agriculture officer, provincial finance officer, health promotion officer, district agriculture officer, and alternative crop program coordinator, all of whom were involved in the alternative crop promotion program. Key informant interviews were conducted in English by the lead author and a trained research assistant in Chipangali using the interview guides. The interview guides pursued questions regarding perspectives on the key factors that affected the success of the alternative crop promotion initiatives, and lessons learned from the program. The full interview guide can be found in Supplementary Guide 1. Interviews were conducted in government offices, restaurants, or homes, near or where key informants were located. Interviews were audio recorded and transcribed verbatim by the lead author. Interview and focus group transcripts were loaded into Atlas.ti for coding and analysis. Data were initially coded according to pre-identified categories derived from the research questions and then reviewed to identify emergent themes and inductively coded into sub-categories. Ethics approval for this study was obtained from University of Zambia Biomedical Research Ethics Committee (UNZABREC) (REF. No. 6011-2024) and National Health Research Authority (NHRAR-R-2083/20/10/2024).

Informed consent was secured from all participants before their involvement in the study. We ensured the confidentiality of the data obtained. Each participant provided written informed consent, facilitated by a trained member of the study team. Potential respondents had the opportunity to ask questions about the research before deciding whether to participate. Those who agreed to participate were interviewed. Participants received written informed consent forms and information sheets detailing the study, its risks and benefits, and emphasizing the protection of confidentiality.

## 3. Results

The success of the alternative crop promotion initiatives was influenced by resource availability, market dynamics, and farmers’ perceptions of tobacco farming. Our findings illustrate the success of the alternative crop promotion program, expressing challenges and lessons learned from the promotion of alternative crop cultivation program in Chipangali district.

### 3.1. Success of Alternative Crop Initiatives

The success of the alternative crop promotion initiatives has been shaped by resource availability, market dynamics, and farmers’ perceptions of tobacco farming. Many farmers emphasized the importance of access to essential inputs such as fertilizers, seeds, and training. Without these, transitioning to alternative crops is challenging. Some farmers noted that growing alternative crops is difficult because of the high cost of fertilizers and they also highlighted that they cannot grow enough crops to sustain themselves without fertilizer. Farmers went on to further emphasize the centrality of essential inputs such as fertilizers and seeds in determining their ability to transition effectively to alternative crops. Without these inputs, many found the shift impractical and unsustainable.

*“Growing alternative crops is difficult due to the high cost of fertilizers. If a company could provide fertilizers and other inputs, even as loans, I would gladly participate”.* (KII Chipangali)

*“Without fertilizers, I cannot grow enough crops to sustain myself. Tobacco may be difficult, but the company gives you everything you need”.* (FGD Chipangali)

*“If the prices for alternative crops on the market could be increased, then I think we could start growing other crops instead of tobacco. Additionally, if there were companies dealing with alternative crops that started giving out loans, then more tobacco farmers might stop growing tobacco”.* (FGD Chipangali)

Furthermore, it was also noted that the financial stability provided by tobacco farming remains a significant factor influencing the success of alternative crop initiatives. Farmers view tobacco as a reliable income source, with established markets and support structures that alternative crops lack. Some participants reported that tobacco farming has a better outcome compared to alternative crops as all items are provided for tobacco farming and it guarantees an income. It was also noted that the financial predictability of tobacco farming is further reinforced by the structured support provided by tobacco companies, including loans, inputs, and access to consistent markets. The absence of such mechanisms in alternative crop initiatives contributes significantly to farmers’ reluctance to switch to alternative crop initiatives.

*“The outcome of tobacco farming is better compared to other crops. You need cash to grow alternatives, but with tobacco, as long as you’re registered with the company, they provide all the necessary items for cultivation”.* (KII Eastern Province)

*“Tobacco is more reliable because it guarantees you money. What other crops offer you that kind of return? I don’t know any”.* (FGD Chipangali)

From the findings it was understood that scepticism about the alternative crop program’s adequacy also impacts success. Farmers expressed disappointment over insufficient meaningful support apart from the lessons.

*“I know many people who joined, and while I’ve been interested, they haven’t received anything meaningful apart from lessons so far. It’s too soon to judge the program’s success”.* (FDG Chipangali)

*“The WHO group came here and told us about alternative crops, but they haven’t provided any real help. The crops they’re promoting aren’t working for me”.* (FDG Chipangali)

“*I heard about the group and their activities, but I was not selected because I grow tobacco. I think they’re meeting to address issues, but honestly, I haven’t seen any real change or development resulting from their efforts*”. (FDG Chipangali)

### 3.2. Challenges and Barriers to Transitioning to Alternative Crops

The findings from the study have highlighted that farmers face numerous challenges in transitioning to alternative crops, including high input costs, dependency on tobacco companies, and market uncertainties. The cost of fertilizers and other inputs was cited as a major barrier. In addition, dependency on tobacco companies made it difficult to transition to alternative crops. Tobacco companies provide inputs, loans, and guaranteed markets, which alternative crop initiatives fail to replicate. This dependency creates a sense of security that alternative crop programs struggle to match.

*“The cost of fertilizers is the biggest challenge. I’m stuck with tobacco because I can’t afford the costs for other crops, even though I’m interested in trying.”* (FGD Chipangali)

*“Tobacco may be stressful, but it seems to pay the bills, right? It’s like that friend who borrows money but always pays you back… sometimes.”* (KII Chipangali)

*“I can’t afford to stop growing tobacco because without the company’s support, I wouldn’t know how to manage without guaranteed input and market.”* (FGD Chipangali)

The study also noted that market uncertainties associated with alternative crops further deter farmers. Unlike tobacco, which has a predictable and reliable market, alternative crops often lack established supply chains, leaving farmers uncertain about profitability. Additionally, some participants also reported that apart from market uncertainties, the guarantee provided by tobacco over alternative crops acts as barrier to transitioning. The participants also cited environmental challenges, such as unpredictable rainfall, contributing to the challenges of switching to alternative crops. It was also pointed out that some farmers fear that alternative crops may not withstand local climatic conditions.

*“I can’t switch to alternative crops if I’m not sure where I will sell them. Tobacco has a market, and I know where to take it every time.”* (FGD Eastern Province)

*“Rainfall here is very unpredictable. I’m not sure if the alternative crops would survive well, so I prefer tobacco for now.”* (FGD Chipangali)

### 3.3. Lessons Learned from the Promotion of Alternative Crop Cultivation

Participants in the study noted that the experiences of the provide valuable lessons for improving the effectiveness of alternative crop initiatives. However, inclusiveness in program design would be very important. Farmers further expressed frustration over the selective nature of current alternative crop initiatives. The importance of making the alternative crop initiatives more inclusive was emphasized, urging program designers to eliminate restrictive eligibility criteria. They suggested that all interested farmers, including those currently engaged in tobacco farming, should be allowed to participate. This inclusivity would ensure broader access to knowledge and benefits, enabling farmers to collectively assess the program’s efficacy.

*“The WFP group should stop being selective and allow anyone to join who is interested. If the program fails, they can fail on their own rather than sidelining others. The alternative crops initiative needs to be open to everyone. Even farmers who are still growing tobacco should be allowed to join. We all need to learn and benefit together.”* (FGD Chipangali)

Additionally, participants pointed out that providing tangible support, such as inputs and financial assistance, would be vital in helping transition to alternative crop initiative. While training is valuable, farmers stressed that it must be complemented by material resources such as seeds and fertilizers to facilitate meaningful participation.

“*If a company could provide fertilizers and other inputs, even as loans, I would gladly participate.”* (FGD Eastern Province)

*“Training is good, but without tangible support like seeds or fertilizers, it’s hard to take the next step. Support must go beyond lessons.”* (KII Eastern Province)

“For me, I can’t stop growing tobacco because it’s the most beneficial crop in this country. I can use a small piece of land to grow tobacco and benefit more than I would using the same size of land for other crops. I wouldn’t benefit as much. Maybe if they provided inputs, some farmers might consider stopping, but for me, I don’t think I’m ready to stop”. (KII Eastern Province)

“*The participants also highlighted that addressing environmental challenges, particularly water scarcity, is important to the success of alternative crop initiatives. Farmers emphasized the need for programs to consider local climatic conditions and to support the cultivation of crops suited to these environments. This includes prioritizing drought-resistant crop varieties capable of withstanding unpredictable rainfall patterns*”. (FGD Chipangali)

*“Alternative crops need to be adapted to the local environment. If the weather is unpredictable, we need drought-resistant varieties or irrigation systems”.* (FGD Chipangali)

*“It would be nice for the program to prioritize drought-resistant crop varieties capable of withstanding unpredictable rainfall patterns to help us farmers.”* (FGD Chipangali)

Additionally, farmers stressed the importance of developing irrigation systems to mitigate water scarcity and sustain agricultural productivity. Infrastructure such as boreholes was identified as a potential solution to support both crop cultivation and livestock needs. By implementing such adaptive measures, alternative crop initiatives can enhance their resilience to environmental challenges and improve their appeal to farmers.

*“If we had boreholes to provide water for our animals and gardens, it could make a significant difference to help transition to alternative crops”. (*KII Eastern Province).

“*Maybe if we can have workshops focusing on drought-resistant crops or how to improve water access, it would help those interested in alternatives while also supporting those farming tobacco*”. (FGD Chipangali)

From the results of the study, it can be seen that alternative crop promotion initiatives have faced significant challenges due to high input costs, market uncertainties, limited support, and dependency on tobacco companies. Despite these barriers, the program’s experiences highlight opportunities for improvement. Inclusive programming that ensures equitable access for all farmers, regardless of their current engagement in tobacco farming, can encourage broader participation and build trust within farming communities. Providing tangible support, such as access to seeds, fertilizers, and tools, alongside financial incentives like loans or subsidies, can reduce the upfront burden of transitioning to alternative crops. Furthermore, addressing environmental challenges such as water scarcity and unpredictable weather patterns through adaptive measures like drought-resistant crop varieties and irrigation systems is important to ensure the sustainability of these initiatives. A farmer-centered approach that integrates these elements is paramount to enabling farmers to shift from tobacco farming to sustainable alternatives. By addressing both the economic and environmental realities that farmers face, future initiatives can create an environment where alternative crops are not just viable but competitive and attractive to rural communities.

## 4. Discussion

Tobacco farming is considered an important driver of development in many countries but it’s also associated with negative health and environmental risks (26). Despite these highlighted negative effects of tobacco, many farmers have continued to cultivate tobacco, and thus alternative crop cultivation initiative programs among tobacco farmers have stalled (27). This qualitative study provides insights into the success, challenges and lessons learned from the alternative crop promotion initiatives in Chipangali District of Zambia. In this current study one of the factors that affected the success of alternative crop initiatives among tobacco farmers was access to essential farming inputs such as fertilizers, seeds, and training. Furthermore, participants expressed that growing alternative crops was difficult because of the high cost of farming inputs and this hindered them from growing enough crops to sustain themselves. The findings were supported by study in Thailand where financial support towards farming found to be critical in facilitating a successful switch to alternative crop farming (28). Similar to the studies in Indonesia, Philippines and Kenya (4, 5, 29) and Malawi (30). The findings emphasize the importance of farming inputs such as fertilizers and seeds in determining tobacco farmers to transitioning to alternative crops.

Supply chain management is another factor that has been shown to influence tobacco cultivation, including transitioning to alternative crops (31). In this study farmers viewed tobacco farming as a reliable source of income, with established markets and support structures that alternative crops lack. Some participants reported that tobacco farming has a better outcome compared to alternative crops as all items are provided for tobacco farming and it guarantees an income. This in line with many studies in tobacco-growing countries that suggest that a robust supply chain of tobacco is one of the main reasons why farmers are hesitant to venture into alternative crop initiative, along with the perceived economic viability and access to markets (32). The results suggest that the financial cost farming inputs and availability of market has a significant impact on the success of the alternative crop initiatives.

The improved tobacco supply chain expressed by most participants could be due to increased use of contract farming. Majority of the tobacco farmers especially in low and middle income countries engage in contract farming with tobacco companies (33, 34). Moreover, In this study, it was also noted that the financial predictability of tobacco farming was reinforced by the structured support provided by tobacco companies, including loans, inputs, and access to consistent markets. The absence of such incentives contributed to farmers’ reluctance to switch to alternative crop initiatives. Farmers felt that even though it was difficult to grow tobacco, companies were available to provide them with farming inputs in form of contracts, findings which are consistent with previous studies conducted in Bangladesh (35), Indonesia and Philippines (4). This is because the tobacco companies guarantee the purchase of the tobacco crops produced by farmers and thus farmers feel secure that their farming efforts will not be in vain. This is further supported by findings in Thailand where tobacco farmers revealed that they were receiving consistent farming support and the market for tobacco was guaranteed through contractual agreements (36). Similarly, in Bangladesh, farmers’ decisions to cultivate tobacco were based on short-term financial support of growing tobacco and better access to credit provided by the tobacco companies compared to other alternative crops (32). The contract arrangements allow farmers to obtain essential inputs such as seeds, fertilisers and pesticides on loan to be paid back on selling their leaf to the lender (23). The findings in this study suggests the need for government and others stakeholder on tobacco control to provide meaningful financial support in form of loans to as to encourage alternative crop farming initiatives.

In this study, skepticism about the alternative crop program’s adequacy also impacted successful transitioning to alternative crops. Farmers expressed disappointment over insufficient meaningful support apart from the lessons. This was affirmed by key informants who stated majority of the farmers who were willing to transition to alternative crop, but no meaningful support was given other than the lessons received during alternative crop initiatives program. Additionally, both farmers and informants felt that the crops they were promoting were not appealing. Echoing the perspectives of farmers in Brazil (37) and Pakistan (38). The findings were further supported by a qualitative study in Kenya where government participants noted that although attempt to guide farmers towards alternative crops were in place, they did not have the resources to support the initiatives and were less able to form meaningful relationships with farmers, and thus had less impact on the decision-making processes of their clients (5). Similarly, in Malawi, the most prominent barrier to diversification efforts identified by respondents was an overwhelming sense of lack of alternatives. It was reported that crop diversification was always on top of agenda of the Malawian government but the implementation was always a challenge despite several advocacy attempts and sensitisation programs (13). The findings indicate the need for increased support by the government to promote alternative crop initiatives among tobacco farmers.

Several challenges hinder the successful transitioning to alternative crops among tobacco farmers (39, 40). In the current study, farmers reported many challenges ranging from high input costs, dependency on tobacco companies, and market uncertainties. The cost of fertilizers and other inputs were cited as a major barrier to transitioning to alternative crops with majority reporting the cost to be too high. These findings mirrored those of Thailand (28), (12) Mozambique and Malawi (3) where high cost of farming inputs and unpredictable markets were cited as major constraints to alternative crop initiatives despite many farmers expressing transition.

In addition, farmers in these FGDs reported that dependency on tobacco companies made it difficult for them to transition to alternative crops. This is because tobacco companies provide the farmers with farming inputs, loans, and guaranteed markets, which alternative crop initiatives fail to replicate. This dependency creates a sense of security that alternative crop programs struggle to match. This findings were in tandem with several studies conducted in Indonesia, Philippines and Bangladesh (4, 32) and Malawi (3). This is due the government’s limited support such as subsidized inputs and extension services which has created a situation where tobacco companies now provide farming input supports on loan, extension services to support crop cultivation thereby tying farmers to these companies ensure continuation of tobacco farming. Findings indicate that improving farmers’ access to farming inputs and low interest loans could lead to the success of the alternative crop programs (41). Because farmers complained consistently about low cost of alternative crops, improving value chains would motivate farmers to shift away from tobacco cultivation (19). Government should, therefore, ensure they support farmers with farming inputs so to motivate them to transition to alternative crop farming.

The perceived lack of the readily available market for alternative crops was a major concern raised by most farmers, who had indicated they were not willing to cultivate other crops. The study also noted that market uncertainties associated with alternative crops further deter farmers. Unlike tobacco, which has a predictable and reliable market, alternative crops often lack established supply chains, leaving farmers uncertain about profitability. This was comparable to a qualitative exploration of farmers’ perspectives in Indonesia and Philippines where uncertainties about readily available market for alternative crops was a major barrier to switching to alternative crops among tobacco farmers (4). This phenomenon of ready market appears to be common theme in several African studies that have examined barrier to alternative crop diversification among tobacco farmers (5, 40, 42). For instance, a study in Uganda revealed that although alternative crops were economically feasible, there were concern about tobacco markets being uncertain (43). This also collaborated well with a survey conducted in three countries including Kenya, Malawi, and Zambia where majority of tobacco farmers expressed lack of a readily available markets as the barrier to successful transition to alternative crop (6). Findings call for the development of institutions that foster value chain development for alternative crops. This confidence of readily available markets could be due to the structured supply chain and corresponding benefits of contractual relationships with tobacco companies. This is also because most finance institutions view small scale tobacco farmers as high risk to finance leaving tobacco buying companies as the sole source of financing for farmers (34). The findings suggest that interventions that identify and link buyers who form equitable partnerships with farmers have the potential to motivate tobacco farmers to shift to alternative crops. Therefore, government must ensure a well-developed supply chains for the proposed alternative crops to attract more tobacco farmers to the alternative crop initiatives.

Additionally, some participants also reported that the guarantee provided by tobacco over alternative crops acts as barrier to transitioning. The participants cited environmental challenges, such as unpredictable rainfall, contributing to the challenges of switching to alternative crops. It was also pointed out that some farmers fear that alternative crops may not withstand local climatic conditions. These findings conquered those of the studies in Indonesia and South India where tobacco farmers viewed tobacco as outstanding plant able to endure the harsh weather conditions, such as low rainfall and poor soil conditions than other alternative crops (6, 39, 44). The findings suggest the need to priotise alternative crops that are to withstand the weather conditions of the local setting in order encourage tobacco farmers to transition.

Participants in the study noted that the experiences of the program provide valuable lessons for improving the effectiveness of alternative crop initiatives. However, inclusiveness in program design would be very important. Farmers further expressed frustration over the selective nature of current alternative crop initiatives. The importance of making the alternative crop initiatives more inclusive was emphasized, urging program designers to eliminate restrictive eligibility criteria. They suggested that all interested farmers, including those currently engaged in tobacco farming, should be allowed to participate. This inclusivity would ensure broader access to knowledge and benefits, enabling farmers to collectively assess the program’s efficacy. Similar concerns were raised in the study conducted in Kenya where agricultural officers said it was important for cooperating partners to ensure that the deserving famers were helped unlike giving relatives or close allies as beneficiaries (5). The findings suggest that if the government invest in other crops, there are possibilities for viability of acceptability among tobacco farmers. For instance, in Malawi, tobacco famers expressed common sentiments that if there were alternatives to tobacco, there would be no opposition to pursuing them by the government with some calling for World Health Organization to present alternatives to that would complement the initiative (3).

Additionally, participants pointed out that providing tangible support, such as inputs and financial assistance, would be vital in helping transition to alternative crop initiative. While training is valuable, farmers stressed that it must be complemented by farming inputs such as seeds and fertilizers to facilitate meaningful participation. Similarly, key informants also gave the similar sentiments. The findings affirms the finding in Indonesia that demonstrated that access to farming inputs or credit could play a substantial roles in tobacco farmers’ economic decision making to transition to other crops (39). Similar findings were noted in Bangladesh (45–47). Therefore, to shift tobacco farmers to other alternative crops, the Government should increase the farmers’ educational level and offer inputs, loans and other incentives to promote alternate crops initiatives.

In this study, participants also highlighted that addressing environmental challenges, particularly water scarcity was important to the success of alternative crop initiatives. Farmers emphasized the need for programs to consider local climatic conditions and to support the cultivation of crops suited to these environments. Similar sentiments were shared by tobacco farmers in a Tanzanian study (48). The study findings serve as basis for the governments to pursue alternative crop initiatives. However, the key to mitigating these challenges lies in prioritizing drought-resistant crop varieties capable of withstanding unpredictable rainfall patterns. The finding suggest that the farmers had acquired important lessons from the alternative crop initiative sensitisation programs.

Additionally, farmers in this study stressed the importance of developing irrigation systems to mitigate water scarcity and sustain agricultural productivity. Infrastructure such as boreholes was identified as a potential solution to support both crop cultivation. Similar perspectives were noted in India and Malawi (49, 50). Finding from this study indicate that tobacco farming is profitable because of the support of the extended towards its cultivation by contract companies. In the similar manner the cultivation of other alternative crops should be left to the providence of natural factors. Hence shifting from farmers to alternative crop farming will only be possible only if similar treatment on par with tobacco is given to selected alternative crops expected to get comparable returns.

This study has some limitations. The use of purposive sampling and small sample size, restricts the generalisability of findings to the entire district of Chipangali and the country at large. Although the FGDs were used in this study to ensure that all participants expressed their perspectives on alternative crop initiatives. The follow-up FGDs may have provided detailed information and clarified some of the perspectives established during data analysis. Furthermore, there is possibility that the meaning of some of the important aspects of the interview guide were lost during translation from the local language to English. However, to account for this, an assistant researcher in the study are fluent in the local language was involved in the study. However, this study has provided important baseline data on the success, challenges and lessons from the implementation of the alternative crop promotion program among tobacco farmers which could help the program to succeed.

## 5. Conclusion

This study has shown that the alternative crop promotion initiatives in Chipangali faced several challenges including high farming input costs, market uncertainties, limited support, and dependency on tobacco companies. Inclusive programming that ensures equitable access for all farmers, regardless of their current engagement in tobacco farming, could encourage broader participation and build trust within farming communities. Providing tangible support, such as access to seeds, fertilizers, and tools, alongside financial incentives like loans or subsidies, can reduce the upfront burden of transitioning to alternative crops. Furthermore, addressing environmental challenges such as water scarcity and unpredictable weather patterns through adaptive measures like drought-resistant crop varieties and irrigation systems was viewed important by participants to ensure the sustainability of the initiatives. It is important that facilitators of alternative crop programs are addressed by tobacco farmers to ensure successful implementation of alternative crop promotion programs. In doing so, a farmer-centered approach that integrates all these elements raised would be paramount to enabling farmers to shift from tobacco farming to sustainable alternative crops.

## Author Contributions

Conceptualization, M.M. (Mercy Mataliro) and C.Z ; methodology, C.Z, and M.M. (Mercy Mataliro); formal analysis, (Mercy Mataliro); investigation, (Mercy Mataliro) ; writing—original draft preparation, (Mercy Mataliro); writing—review and editing, C.Z, M.M (Martha Mutalange) and M.M. (Mercy Mataliro) ; supervision, CZ and M.M (Martha Mutalange) ; revisions, C.Z, M.M (Martha Mutalange) and M.M. (Mercy Mataliro); All authors have read and agreed to the published version of the manuscript

## Data Availability

The data supporting the findings of this study are available from the corresponding author upon reasonable request. For any additional information or queries regarding the data, please contact.

## Acknowledgments

The authors would like to acknowledge the participants for taking their time and valuable information, without them which this study couldn’t be successful. The authors would also like to acknowledge Professor F Goma, Agatha Shula, Chipangali District Health Management and Chipangali District Agriculture officers.

